# Advanced electrocardiography predicts cardiac involvement and incident arrhythmias in Fabry disease

**DOI:** 10.1101/2025.03.02.25323189

**Authors:** Ravi Vijapurapu MBChB, Maren Maanja, James Every, Todd Schlegel, Ashwin Roy, James C Moon, Derralynn A Hughes, Michel Tchan, Tarekegn Geberhiwot, Martin Ugander, Richard P Steeds, Rebecca Kozor

## Abstract

**Background:** Fabry disease (FD) is an X-linked disorder with progressive myocardial sphingolipid deposition, causing premature cardiovascular morbidity and mortality. Advanced electrocardiography (A-ECG) has the potential to predict cardiac involvement.

**Objectives:** To evaluate the predictive power of A-ECG in identifying: 1) early cardiac involvement defined as low myocardial T1 on cardiovascular magnetic resonance (CMR), 2) adverse cardiovascular outcomes, and 3) heart age.

**Methods:** In this longitudinal multi-centre study, patients underwent same-day CMR and digital ECG, analysed using in-house software, including conventional ECG, derived vectorcardiographic, and singular value decomposition measures of waveform complexity parameters. Significant A-ECG variables were identified using stepwise forward regression and incorporated in a multivariable logistic regression A-ECG score. A Youden index was applied to identify best threshold score and bootstrapping performed to calculate the area under the receiver operating characteristics curve (AUC), sensitivity, specificity, and 95% confidence intervals (CI).

**Results:** In 155 patients (40% male, age 46±14 years, 39% on enzyme replacement therapy), left ventricular mass was higher in males compared to females (106 vs. 59 g/m^2^, p<0.001), 80% had low native T1, and 51% (70/136) had late gadolinium enhancement. A-ECG heart age was higher than chronological age in all patients (57±20 vs. 46±14 years, p<0.001). The heart age gap was strongly associated with T1 lowering and progressive LVH. Multivariable A-ECG scores for detecting low T1 had an AUC [95%CI] of 0.82 [0.75-0.89], sensitivity 72 [55-95] %, and specificity 85 [66-71] %; any arrhythmia 0.89 [0.82-0.95], 82 [68-94] %, 88 [70-96] %; or atrial fibrillation 0.89 [0.80-0.96], 92 [77-100] %, 83 [76-92] %, respectively. No predictors of heart failure hospitalisation or mortality were found.

**Conclusion:** A-ECG analysis has good diagnostic performance for predicting low native T1 and the occurrence of arrhythmias in Fabry disease but not for heart failure hospitalisation or death.

## Introduction

Fabry disease (FD) is an X-linked lysosomal storage disorder, in which a deficiency in the enzyme alpha-galactosidase A leads to gradual accumulation of sphingolipids in multiple tissues, including the heart, kidneys and brain (1, 2). Cardiovascular magnetic resonance imaging (CMR) with native T1 mapping can identify myocardial sphingolipid deposition.

Cardiovascular disease is the leading cause of morbidity and mortality in Fabry disease (3). Cardiac involvement, such as progressive left ventricular hypertrophy (LVH) (4), myocardial fibrosis, congestive cardiac failure, arrhythmia and sudden death (5), occurs in up to 78% of patients (6), with men usually affected earlier than women. Although disease modifying therapies are increasingly used, they are less effective once advanced cardiac disease is present. This suggests that earlier introduction of treatment may be required (7, 8); however, a greater understanding of early markers of disease is needed in order to identify the optimal approach to treatment.

Conduction system involvement is known to occur early in the disease process often prior to the onset of myocardial hypertrophy (6), however, there are no ECG variables that are pathognomonic to Fabry disease. Advanced ECG analysis (A-ECG) is a simple non-invasive technique that analyses a standard resting digital 12-lead ECG, and in other disease processes has been shown to provide superior diagnostic and prognostic information (9). Combining this novel technology with CMR T1 mapping offer the opportunity for reliable detection of early cardiac involvement that have the potential to act as an alternate trigger for therapy.

Cardiovascular disease is the leading contributor to morbidity and mortality in Fabry, with insidious progression of sphingolipid accumulation from early age. Progression, however, is variable between individuals in the same family, and is not directly linked to genotype. Heart age is a concept developed to reflect biological variability in disease expression in the general population, and advanced age better predicts long-term outcome. Enzyme replacement and oral chaperone therapies are expensive, and early identification of cardiovascular involvement, together with the ability to identify those at greatest risk offers the opportunity of targeting therapy to those most in need.

The aim of this study was to evaluate the predictive power of A-ECG markers in identifying: 1) early cardiac involvement defined as low myocardial T1 on CMR, and 2) adverse cardiovascular outcomes including arrhythmias, and 3) heart age.

## Methods

### Study population

This was a multi-centre study with patients recruited from two groups. The larger group were patients recruited to the prospective, observational international Fabry400 study (NCT03199001) from University Hospital Birmingham, Royal Free Hospital London and the National Hospital for Neurology and Neurosurgery London, and Westmead Hospital Sydney Australia. Patients were eligible for the Fabry400 study if they were ≥ 18 years of age and had a genetically confirmed diagnosis of FD. Patients were excluded if they had an absolute or a relative contra-indication to CMR. A smaller proportion of study patients were taken from consecutive adults with genetically proven FD attending the UHB Rare Diseases Centre. Patients were recruited from August 2016 to November 2019. Local ethics approval was obtained for the Fabry400 study (14/LO/1948) and the Health Regulation Authority in the UK waivers consent for the use of data historically collected as part of a clinical service where its inclusion in research is secondary. This study conformed to the principles of the Helsinki Declaration.

### CMR imaging

All participants underwent CMR at 1.5 Tesla (Avanto (UK), Aera (Australia); Siemens Healthcare, Erlangen, Germany) using a standard protocol including LV cines in short axis (SAX), 4-chamber, 2-chamber and 3-chamber views. Native T1 mapping was performed pre-contrast on basal and mid left ventricular SAX slices using a modified Look-Locker inversion recovery (MOLLI) sequence. Late Gadolinium enhancement (LGE) imaging was performed using phase sensitive inversion recovery (bolus administration of gadolinium 0.1 mmol/kg body weight, Gadoterate meglumine, Dotarem, Guerbet S.A., France)

### CMR analysis

All images were analysed using CVI42 software (Circle Cardiovascular Imaging Inc., Calgary, Canada). Cardiac chamber volumes and LV mass (LVM) (papillary muscles included in mass) were quantified on all subjects from a pre-contrast breath-held SAX stack of balanced steady-state free precession cine images, using previously described manual contouring methodologies (10). Left ventricular hypertrophy (LVH) was defined as increased indexed LVM on CMR according to age and sex matched normal reference ranges (males: 74±8.5g/m^2^ and females: 63±7.5g/m^2^) (11). Pre-contrast native T1 was evaluated in the basal and mid LV using a region of interest technique, as previously described (12–14). Normal ranges were defined as the mean ± two standard deviations based on study centre specific healthy controls (London: males mean 956±27ms, lower limit 902ms; females mean 978±34ms, lower limit 910ms. Birmingham: males mean 947±28ms, lower limit 890ms; females mean 958±30ms, lower limit 898ms. Sydney: males mean 947±24ms, lower limit 893ms; females mean 965ms±31ms, lower limit 903ms). Native T1 was subsequently categorised as low or normal.

### Advanced ECG analysis

Standard resting 12-lead ECGs (10-second recordings, sample rate 250Hz) were captured using local ECG machines in all participants and subsequently exported and stored as digital .xml files.

A-ECG semi-automatic software developed in-house was used to analyse the xml files, and this encompassed conventional ECG measures of durations and amplitudes, derived vectocardiographic (VCG) measures, and singular value decomposition measures of waveform complexity.

Additionally, A-ECG Heart Age was calculated by inputting the following parameters into a predefined mathematical formula, as described by Lindow et al (18).

1. Conventional ECG parameters: heart rate, R-R interval, P wave, PR interval, QRS duration, QT/QTc, Tq interval, cardiac axis and electrical amplitudes on ECG.
2. Vectorcardiographic data: spatial QRST angle (mean and peak), spatial ventricular gradient and QRS/T wave components, spatial QRS and T wave axes, waveform amplitudes/areas, spatial QRS and T wave velocities.
3. QRS and T wave complexity based on SVD.

The A-ECG heart age is an estimate of a person’s age based on their heart, and can be compared to their chronological age, with the difference being ’heart age gap’.

### Statistical analysis

Statistical analyses were carried out using SPSS 22 (IBM, Armonk, NY) and R version 3.4.3 (R Foundation for Statistical Computing, Vienna, Austria). Normality was checked using the Shapiro-Wilk test and by visual review of histogram data for all variables. Normally distributed data were expressed as mean ± standard deviation and compared using the independent-samples t-test or one-way ANOVA with post-hoc Tukey correction. Non-normally distributed data were described using median and IQR with comparisons between groups using Mann-Whitney U or Kruskall-Wallis test, as appropriate. Categorical data were described as frequencies or percentages and chi-squared testing utilised when comparing proportions of a variable between two groups.

Diagnostic and prognostic scores for outcome markers were determined using continuous A-ECG data and stepwise forward logistic regression. A maximum of one parameter for ten events was used. To confirm accuracy of the scores derived from regression modelling the area under the ROC curve (AUC) was calculated and bootstrapped 2000 times to obtain 95% confidence intervals (95% CI) and the score with the higher AUC was used. A Youden index was utilised to identify the cut off scores that optimised sensitivity and specificity. A p-value of <0.05 was considered statistically significant.

## Results

### Participant characteristics

In total, 155 Fabry patients were included in this study, with a mean age of 46±14 years. There were 62 men (40%) and 93 women (60%). A non-classical cardiac mutation was present in 30% of patients and 61 patients (39%) were on ERT at time of recruitment. CKD stage 3 or above was present in 8% of the total cohort with a lower eGFR in men (87 [59-90] vs. 90 [79-90] ml/min/1.73m^2^, p=0.040). Ten percent of patients had a confirmed diagnosis of hypertension on treatment, but none had known/evidence of ischaemic heart disease at recruitment. Demographic data can be seen in *Table 1*.

**Table 1.**
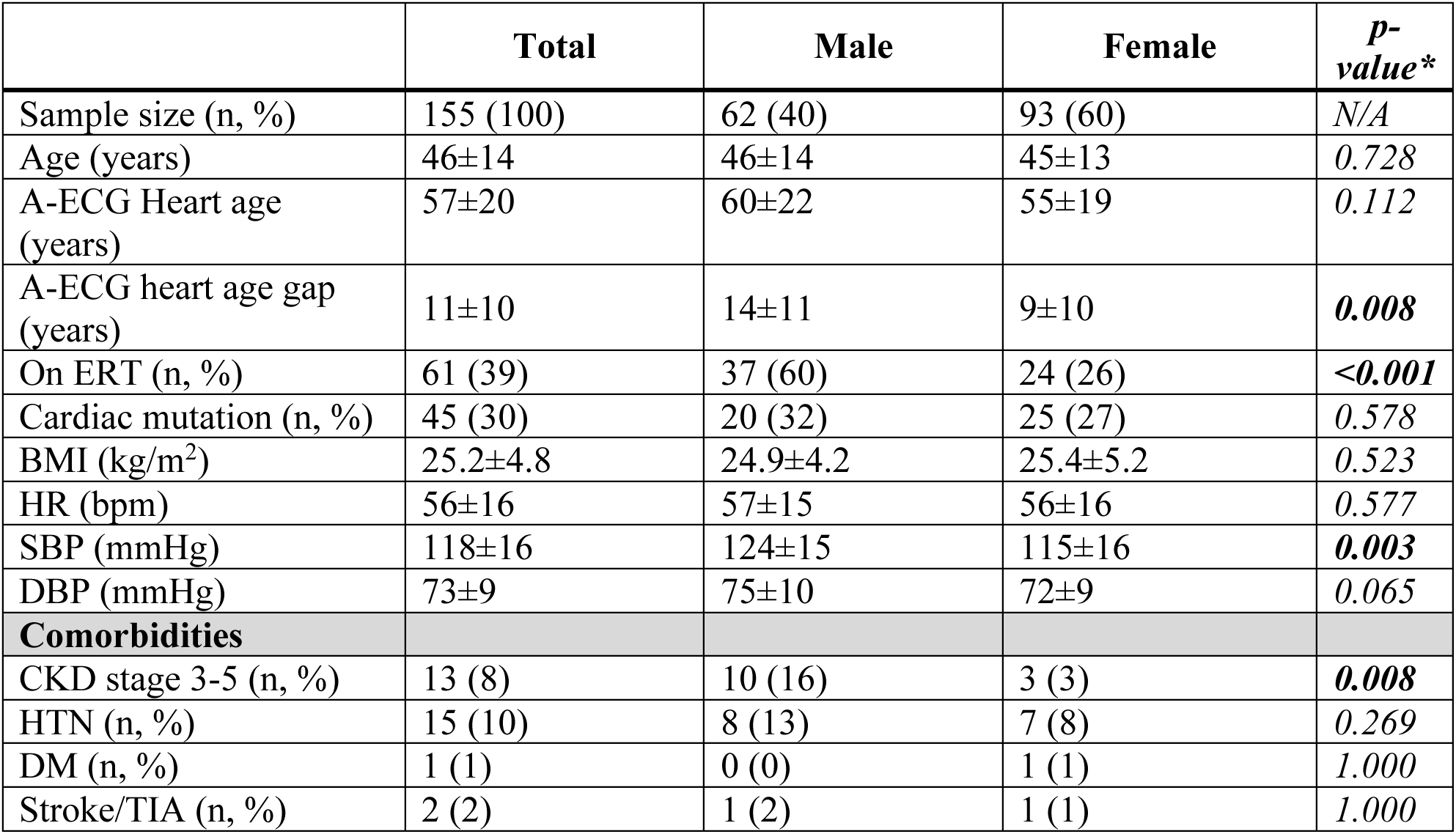
Demographic data for the study cohort.

*Table 2* demonstrates the CMR and blood biomarker data of the study cohort. Of the total cohort, 36% (56/155) had LVH (“LVH positive”) and 64% (99/155) had no LVH (“LVH negative”). There were differences in LVH between the sexes: LVH positive: men – 73%, 41/56; women – 27%, 15/56; LVH negative: men – 21%, 21/99; women – 79%, 78/99. Men had higher LVMi and maximum wall thickness (MWT) compared to women (LVMi: 106.0 vs. 58.8 g/m^2^, MWT: 14.0 vs. 9.0 mm, p<0.001 for both). Of the total cohort, 92 patients (80%) had low native T1 according to local reference ranges: men 40/62, 65% vs. women 52/94, 55%, p=0.004. When split by presence or absence of LVH, 71% had low T1 in the LVH positive cohort (men: 28/41, 68% vs. women 12/15, 80%; p=0.513). In those without LVH 53% had low T1 (men: 12/21, 57% vs. women: 40/78, 51%; p=0.806). Of the total cohort 51% had LGE (defined as visual LGE in two or more segments of the LV). Presence of LGE was more common in men than women (37/54, 69% vs. 33/82, 40%, respectively, p=0.004). Of the LVH positive patients 42/48 (88%) had LGE, compared with 28/88 (32%) in the LVH negative group (p<0.001). High-sensitive troponin and NT-pro BNP were mostly within the normal range, but levels were greater in men.

**Table 2.** Cardiac investigation and biomarker data.

|  | **Total** | **Male** | **Female** | ***P*-value\*** |
| --- | --- | --- | --- | --- |
| **CMR** |  |  |  |  |
| LVMi (g/m<sup>2</sup>)* | 70.0 (57.3-99.8) | 106.0 (77.8-138.4) | 58.8 (53.4-74.1) | **<0.001** |
| LVH positive (n, %) | 56 (36%) | 41 (66%) | 15 (16%) | **<0.001** |
| MWT (mm) | 10.0 (8.3-14.0) | 14.0 (11.8-18.0) | 9.0 (8.0-11.0) | **<0.001** |
| LVEDVi (ml/m<sup>2</sup>) | 72.9±12.9 | 77.5±14.9 | 69.9±10.4 | **0.001** |
| LVESVi (ml/m<sup>2</sup>) | 20.0±7.2 | 21.9±7.6 | 18.8±6.7 | **0.010** |
| LVSVi (ml/m<sup>2</sup>) | 56.4±40.0 | 55.8±12.0 | 56.9 | 0.874 |
| Native T1 – mid LV cavity (ms)* | 944.4 (907.6 – 983.7) | 932.8 (891.1-950.4) | 967.6 (911.0-990.6) | 0.055 |
| Low T1 (n, %) | 92 (80) | 40 (65) | 52 (56) | 0.245 |
| LGE (n, %) | 70/136 (51) | 37/54 (69) | 33/82 (40) | **0.004** |
| RVEDVi (ml/m<sup>2</sup>) | 75.7±15.6 | 78.2±17.8 | 74.2±14.1 | 0.304 |
| RVESVi (ml/m<sup>2</sup>) | 31.2±12.2 | 34.8±16.9 | 29.4±8.9 | 0.241 |
| RVSVi (ml/m<sup>2</sup>) | 46.6±10.0 | 46.3±11.7 | 46.7±9.3 | 0.882 |
| **Biomarkers** |  |  |  |  |
| High sensitive troponin T (ng/L)* | 12.0 (1.0-33.0) | 20.6 (4.2-58.3) | 3.5 (1.0-22.5) | **<0.001** |
| NT-pro BNP (ng/l)* | 12.5 (3.6-47.5) | 18 (2.7-120.3) | 11 (3.9-37.8) | 0.283 |
| eGFR (ml/min/1.73m<sup>2</sup>)* | 90 (74.8-90) | 87 (59-90) | 90 (79.2-90) | **0.040** |

\* non-parametric data so presented as median (IQR)

### A-ECG Heart Age

The mean A-ECG derived heart age across the total cohort was 57±20 years compared to their chronological mean age of 46±14 years, p<0.001. This equates to a “heart age gap” of 11 years (this term refers to the difference between A-ECG and chronological heart ages), indicating patients with Fabry disease had a higher A-ECG heart age compared to chronological age. This was also evident when split by sex – men: 60±22 vs 46±14, p<0.001, women: 55±19 vs 45±13, p<0.001). *Table 3* demonstrates the heart age data in various sub-groups. The A-ECG heart age gap was higher in those patients with LVH (present data here just on LVH positive versus negative). And was higher in patients with low T1 (present data here on low T1 versus normal T1). We then looked at those patients with early disease (no LVH and either low T1 or normal T1) versus established disease with LVH, and heart age gap was significantly higher only in the LVH positive group: (LVH positive: 18±10 years vs. LVH negative/low T1: 8±9 years vs. LVH negative/normal T1 7±8 years, p<0.001).

**Table 3.**
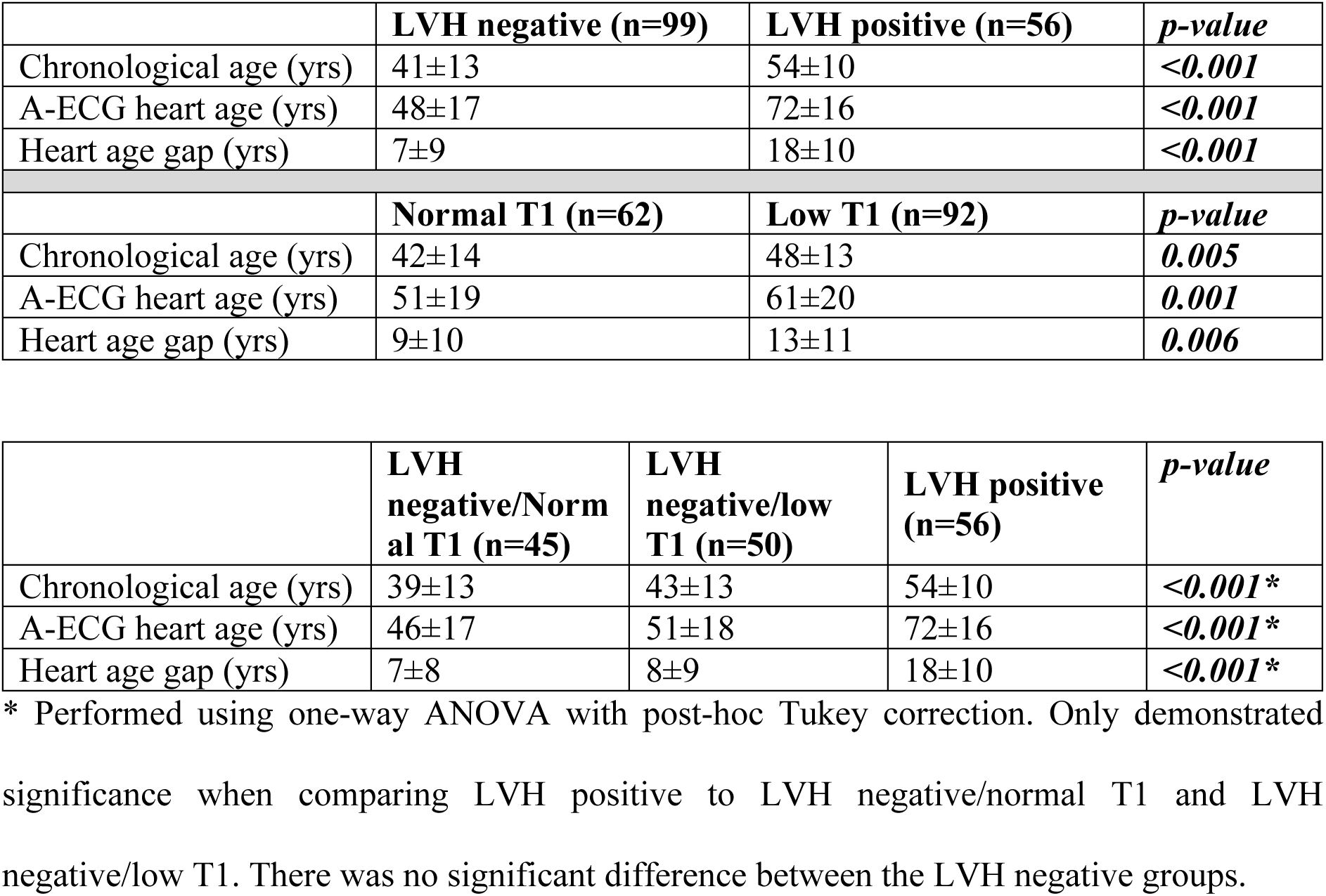
Heart age data

The heart age gap demonstrated a positive correlation with markers associated with advanced cardiac disease. These included the occurrence of an arrhythmia (r=0.293, p<0.05), AF (r=0.276, p<0.01), increasing indexed LV mass (r=0.450, p<0.001, *Figure 2*), the presence of LVH (r=0.471, p<0.01) and low native T1 (r=-0.275, p=0.02, *Figure 3*). The presence of CKD (stage 3 or higher) was also associated with an increasing heart age gap (r=0.209, p<0.05).

**Figure 1.**
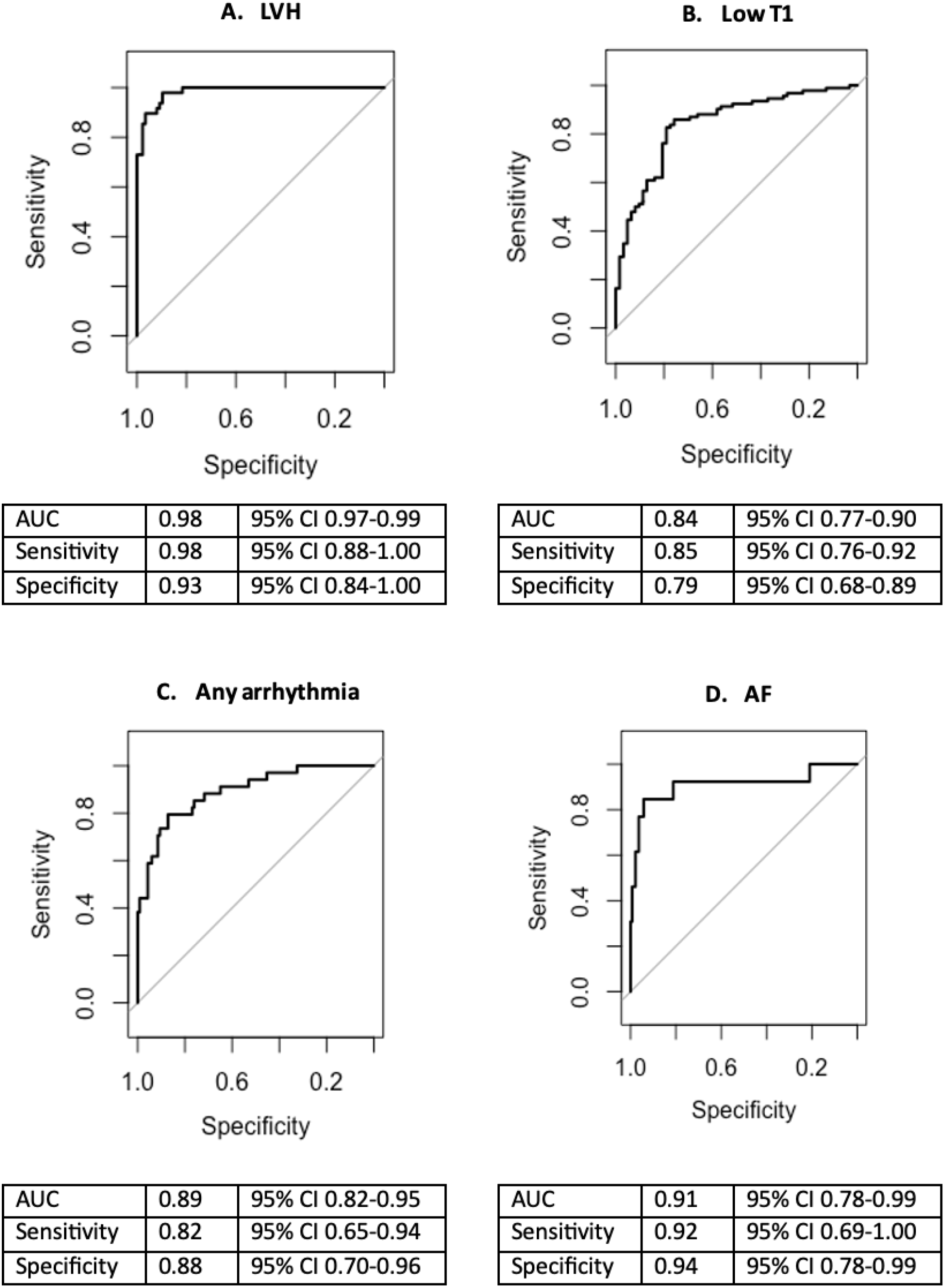
Receiver operating curves demonstrating the sensitivity and specificity of the A-ECG Fabry score.

**Figure 2.**
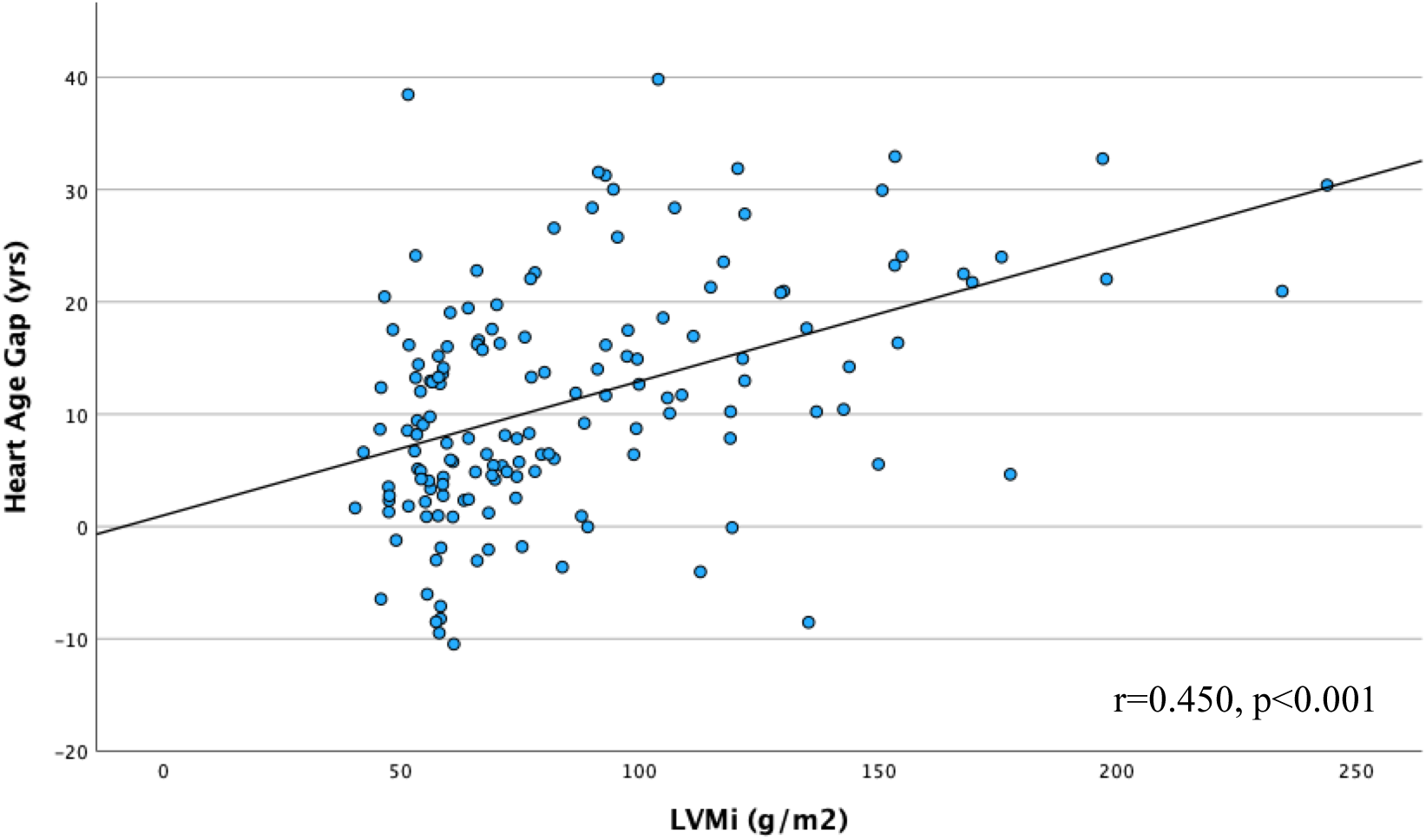
Correlation between heart age gap and LVMi.

**Figure 3.**
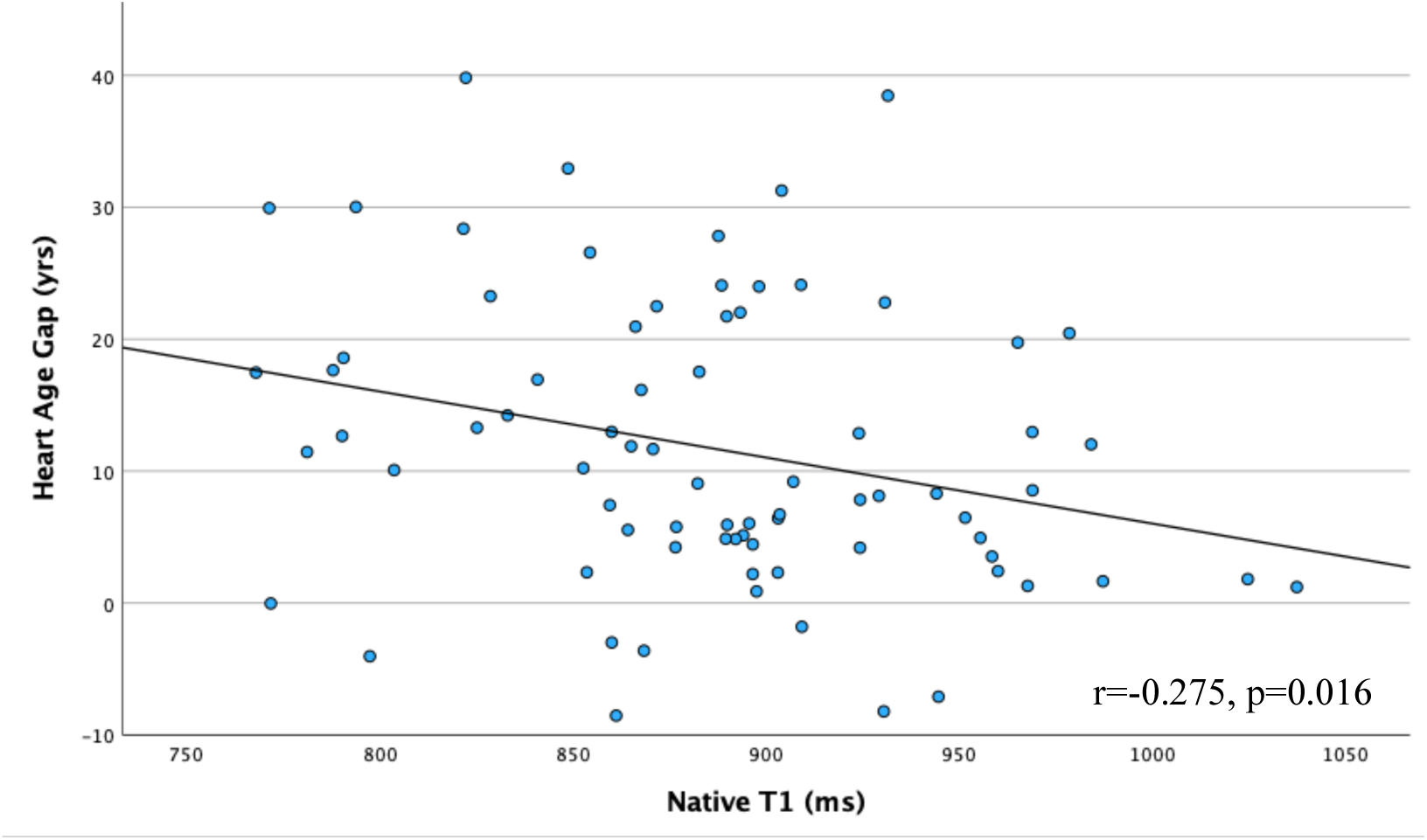
Correlation between heart age gap and native T1

### A-ECG predictors of outcome variables

#### 1. LVH

As described earlier, 36% of the total cohort (n=56) had evidence of LVH as defined by CMR evaluation (66% in men and 16% in women). *Table 4* describes the A-ECG parameters that were found to be predictive for the presence of LVH in FD. This predictive scoring model was found to have an area under the curve (AUC) of 0.98 (95% CI 0.97-0.99) with a sensitivity of 98% (95% CI 88-100) and a specificity of 93% (95% CI 84-100). This can also be seen in *Figure 1a*.

**Table 4.**
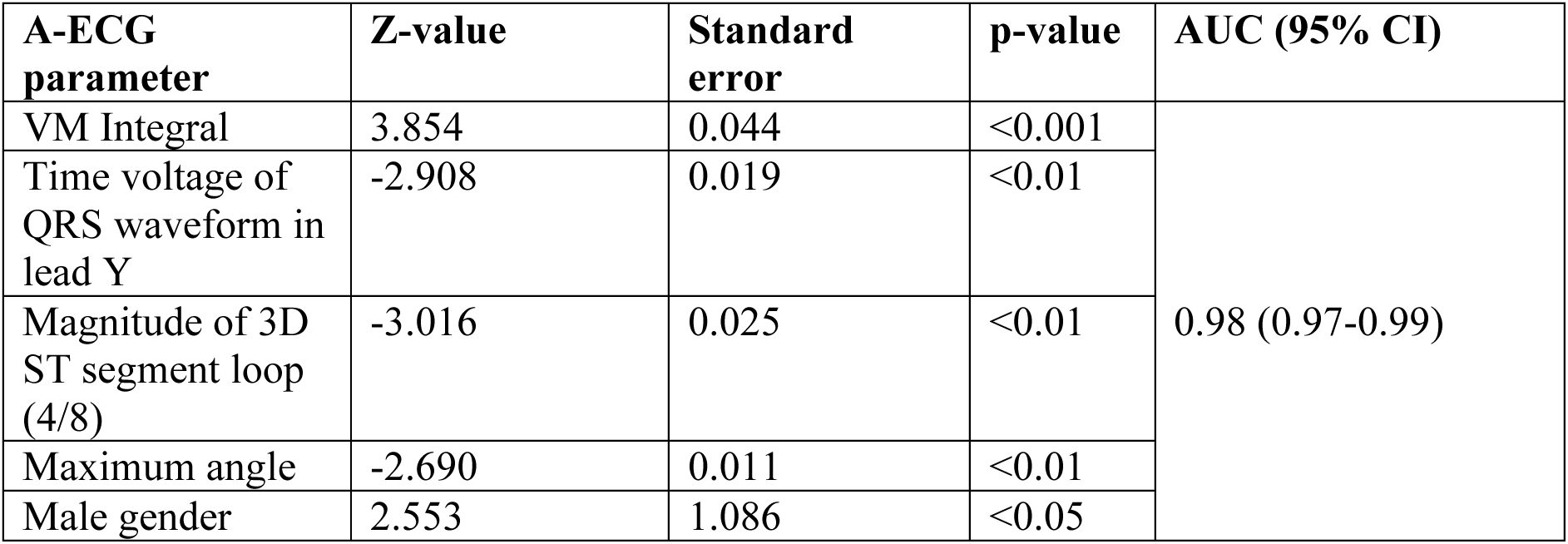
A-ECG parameters predictive of LVH.

#### 2. Low Native T1

Of the total cohort 92 patients (80%) were found to have low T1, with 40 men and 52 women. 50 patients (53%) with a normal LV mass had low native T1, suggestive of “early disease” subclinical sphingolipid deposition (mean native T1: 904±39 ms; men: 12 (13%), 897±27 ms; women 38 (40%), 906±42 ms, p=0.700). *Table 5* demonstrates the individual A-ECG parameters that in combination were found to be predictive of low native T1. This diagnostic score was found to have an AUC of 0.84 (95% CI 0.77-0.90), a sensitivity of 85% (95% CI 76-92) and a specificity of 79% (95% CI 68-89), *Figure 1b*.

**Table 5.** A-ECG parameters predictive of low native T1.

| **A-ECG parameter** | **Z-value** | **Standard error** | **p-value** | **AUC (95% CI)** |
| --- | --- | --- | --- | --- |
| QRS axis | -2.956 | 0.0039 | <0.01 | 0.84 (0.77-0.90) |
| Elevation angle in 3D QRS loop (8/8) | -3.451 | 0.0050 | <0.001 |  |
| Ratio of rightward QRS loop to leftward QRS loop in horizontal plane | 3.078 | 0.0052 | <0.01 |  |
| Elevation angle of the polar vector | -3.565 | 0.0054 | <0.001 |  |
| Magnitude of the ST loop (3/8) | -3.163 | 0.0049 | <0.01 |  |
| Azimuth angle of the 3D QRS loop (6/8) | -3.310 | 0.0051 | <0.001 |  |
| Summed 12 lead voltage | -2.675 | 0.0053 | <0.01 |  |
| Magnitude of 3D QRS loop (3/8) | 1.985 | 0.0050 | <0.05 |  |

#### 3. Arrhythmia

Of the total cohort 34 patients (22%) had an arrhythmic event documented over a mean follow-up period of 3.2±0.6 years. This comprised of 13 patients with AF needing anticoagulation (6 women and 7 men), 12 with atrio-ventricular nodal re-entrant tachycardia (AVNRT) requiring medical therapy (6 women and 6 men) and 13 with non-sustained ventricular tachycardia (NSVT) (6 women and 7 men). No patients had sustained ventricular tachycardia (VT). The mean age of patients with any arrhythmia was 54±10 years (men 53±10 and women 56±9 years).

A combined predictive score for the occurrence of any arrhythmia had an AUC of 0.89 (95% CI 0.82-0.95), sensitivity of 82% (95% CI 65-94) and specificity of 88% (95% CI 70-96). When evaluating predictive markers for AF requiring anticoagulation the predictive score had an AUC of 0.91 (95% CI 0.78-0.99), sensitivity of 92% (95% CI 69-100) and a specificity of 94% (95% CI 78-99). No significant A-ECG predictors were found for the occurrence of NSVT and AVNRT. *Table 6* and *Figures 1b and 2c* describe the individual components of the A-ECG predictive score for any arrhythmia and AF.

**Table 6.** A-ECG parameters predictive for the occurrence of any arrhythmia and AF.

| A-ECG parameter | Z-value | Standard error | p-value | AUC (95% CI) |
| --- | --- | --- | --- | --- |
| 1) Any arrhythmia requiring therapy |  |  |  |  |
| Maximum voltage of Frank Z-lead QRS complex | 4.108 | 0.012 | <0.01 | 0.89 (0.82-0.95) |
| QRS axis | -3.515 | 0.004 | <0.001 |  |
| Amplitude of second singular value of the T wave | 2.873 | 0.006 | <0.01 |  |
| Magnitude of the QRS loop 7/8 | 3.139 | 0.008 | <0.01 |  |
| Maximum spatial velocity of the magnitude of the spatial T wave | -2.556 | 0.008 | <0.05 |  |
| Total QRS loop in frontal plane | -2.527 | 0.011 | <0.05 |  |
| 2) AF requiring anticoagulation |  |  |  |  |
| Maximum voltage of the Z-lead in the QRS complex | 3.55 | 0.015 | <0.001 | 0.91 (0.78-0.99) |
| Absolute area subtended by the T wave in the Z-lead | -2.59 | 0.008 | <0.01 |  |
| Difference between frontal T wave elevation and T wave azimuth | -2.10 | 0.009 | <0.05 |  |
| T Eigenvector 5 | -1.992 | 0.009 | <0.05 |  |

#### 4. Scar burden on CMR

Of those who underwent LGE imaging 70 patients had evidence of LGE (women: 33/82, 40% and men: 37/54, 69%; p=0.004). Those with RV insertion point LGE were not included in data analysis. The components of the score predictive of LGE are shown in *Table 7* (AUC 0.94 [95% CI 0.89-0.97], specificity 91% [95% CI 79-100], sensitivity 86% [95% CI 71-96]).

**Table 7.** A-ECG parameters predictive of LGE

| **A-ECG parameter** | **Z-value** | **Standard error** | **p-value** | **AUC (95% CI)** |
| --- | --- | --- | --- | --- |
| T wave morphology | 4.269 | 0.238 | <0.001 | 0.94 (0.89-0.97) |
| Elevation angle of the spatial ventricular gradient (left sagittal plane) | -3.615 | 0.007 | <0.001 |  |
| Q wave eigenvector 8 | -3.321 | 0.009 | <0.001 |  |
| Absolute area of the T wave in lead Y | 3.152 | 0.007 | <0.01 |  |
| ST Elv 1.8 | -3.135 | 0.007 | <0.01 |  |
| Magnitude of 3D QRS loop (7/8) | -2.772 | 0.007 | <0.01 |  |
| Magnitude of 3D QRS loop (4/8) | -2.173 | 0.007 | <0.05 |  |

#### 5. Heart failure hospitalisation and sudden death

Six patients (3 women and 3 men) required hospitalisation for treatment of decompensated heart failure (HF) and no deaths were recorded during the follow-up period. No significant A-ECG predictors were found for hospitalisation of HF.

## Discussion

This is the first study evaluating the diagnostic and predictive power of A-ECG in Fabry. Detection of early cardiovascular disease with CMR T1 mapping is well recognised and has revolutionised the diagnostic pathway of Fabry cardiomyopathy. This study has demonstrated that A-ECG analysis of the standard 12-lead ECG can be strongly predictive of low T1 and therefore subclinical cardiac disease. From a total of 234 A-ECG variables, a global predictive model incorporating seven of these A-ECG parameters, primarily derived from the QRS and ST vectors, were found in combination to be highly predictive of low T1, with an AUC of 0.84. In addition to its diagnostic power this study has demonstrated the value of A-ECG in predicting adverse arrhythmic outcomes. In particular two A-ECG parameters were demonstrated to be strongly predictive of AF requiring anticoagulation, over a relatively short follow-up time. This study has also shown that A-ECG heart age gap is strongly associated with progressive Fabry cardiomyopathy, with a significantly greater heart age gap in those with LVH compared to those without. Progressive sphingolipid deposition was also associated with an increasing heart age gap.

Abnormalities in the resting 12-lead ECG have historically been described in Fabry cardiomyopathy with more significant electrical changes observed with advancing cardiac disease (19, 20). More recent literature has evaluated the ECG in greater detail and described evidence of electrical abnormality much earlier in the disease phenotype. Nordin et al (12) demonstrated that ECG abnormalities tended to co-segregate with low T1 even in the LVH negative population, suggesting that a combined assessment of ECG and CMR may be beneficial. This was further evaluated by Augusto et al (21), who identified that certain ECG abnormalities within the T wave occurred in a very early pre-phenotypic stage of Fabry cardiomyopathy (LVH negative, normal T1) compared to healthy controls, suggesting that more advanced ECG abnormalities occur much early than previously thought. The results described in this chapter highlight that using more sophisticated A-ECG analysis techniques, a global predictive score of seven AECG parameters predominantly from the QRS and T wave vectors is highly predictive of early cardiac disease.

### Adverse outcome prediction

The use of AI derived advanced ECG analysis as a predictor of arrhythmia has been widely described over the past few years. A study by Christopoulos et al (22) of 1936 healthy volunteers aged between 70-89 years utilised an AI enabled convolutional neural network ECG algorithm to determine the probability of developing AF. They found that using AI-ECG analysis software a predictive model output greater than 0.5 led to a 2-year cumulative risk of AF of 21.5% and a 10-year risk 52.2%, highlighting the benefit of ECG based predictive scoring (22, 23). This methodology however, was slightly more time-consuming requiring specialist computer technology to perform analysis. More recently Sanz-Garcia et al (24) demonstrated the potential use of automated ECG analysis techniques as a low-cost screening strategy for AF in healthy volunteers. They identified 32 specific ECG variables that demonstrated predictive value of AF, with an AUC of 0.776. The A-ECG techniques described in this chapter were similar in simplicity to achieve but yielded a greater number of vectorcardiographic parameters. Despite shorter follow-up and consequently lower number of outcome events, two specific parameters predictive of AF in Fabry were identified with an AUC 0.89, thus highlighting this methodology as potentially valuable predictor of adverse arrhythmic outcomes.

The importance of the QRS-T angle in predicting adverse clinical outcomes has also been described in the literature. A study of 1915 patients presenting to a tertiary centre with symptoms of breathlessness demonstrated that a wide QRS-T angle was a strong discriminator between a diagnosis of acutely decompensated HF and non-cardiac causes of breathlessness (25). Sub study analysis also identified the QRS-T angle as a strong predictor of 2-year mortality in HF. Due to the shorter follow-up time of our study, fewer HF hospitalisation episodes were identified and consequently no statistically significant variables were identified.

### A-ECG heart age

The concept of heart age has been used to evaluate a patient’s cardiovascular age based on their cardiovascular risk profile. The 12-lead ECG is known to alter both as patient’s age and also in the presence of cardiac disease, with certain parameters predictive of adverse clinical outcomes. Integration of electrocardiographic data into heart age calculation using AI technology has been shown to have a greater sensitivity and specificity for assessment of cardiovascular risk when compared to more traditional cardiovascular risk estimation models (26). Recent work by Lindow et al concluded that ECG heart age could accurately be determined from standard ECG measures taken from a 10 second digital ECG without the use of AI techniques (18), thus making this methodology potentially more clinically applicable. They also identified that the difference between A-ECG heart age and chronological age (termed heart age gap) increases with progressive cardiovascular disease and can be predictive of adverse outcomes. This study has supported this theory with a higher A-ECG heart age compared to chronological age in Fabry cardiomyopathy. A higher A-ECG age was also observed in LVH negative men with low T1, suggesting that cardiovascular risk may be altered much earlier in the disease process prior to the onset of LVH, however further prospective validation is necessary (27).

### Limitations

The limitations of this study include the small size of the study cohort, which made detailed sub-analysis impossible due to very low numbers in individual groups. Furthermore, the follow-up time of this cohort was relatively short, resulting in a low detection rate of outcome markers such as individual arrhythmia, HF hospitalisation and mortality. The addition of a healthy volunteer control group and a Fabry validation cohort would have been of significant value, however, this was out of the scope of this initial study and further follow-on work will provide this data (27).

## Conclusion

A-ECG analysis of the resting 12-lead ECG has good diagnostic performance for predicting left ventricular hypertrophy, low native T1 and the occurrence of arrhythmias in Fabry disease. This supports the use of A-ECG both as a screening tool to diagnose (early) cardiac disease, and for identifying those at risk of adverse arrhythmic outcomes.

## Data Availability

All data produced in the present work are contained in the manuscript.

## Acknowledgements

We would like to acknowledge the contribution of the patients and staff within the Fabry centres included in this study. This study is part of the Fabry400 study, which is funded by an investigator led research grant from Sanofi-Genzyme. All co-authors contributed to data interpretation and subsequent editing of the manuscript.

## References

1. Desnick RJ, Brady R, Barranger J, Collins AJ, Germain DP, Goldman M, et al. Fabry disease, an under-recognized multisystemic disorder: expert recommendations for diagnosis, management, and enzyme replacement therapy. Ann Intern Med. 2003;138(4):338–46.

2. Zarate YA, Hopkin RJ. Fabry’s disease. Lancet. 2008;372(9647):1427–35.

3. Linhart A, Kampmann C, Zamorano JL, Sunder-Plassmann G, Beck M, Mehta A, et al. Cardiac manifestations of Anderson-Fabry disease: results from the international Fabry outcome survey. Eur Heart J. 2007;28(10):1228–35.

4. Nordin S, Kozor R, Bulluck H, Castelletti S, Rosmini S, Abdel-Gadir A, et al. Cardiac Fabry Disease With Late Gadolinium Enhancement Is a Chronic Inflammatory Cardiomyopathy. J Am Coll Cardiol. 2016;68(15):1707–8.

5. Mehta A, Clarke JT, Giugliani R, Elliott P, Linhart A, Beck M, et al. Natural course of Fabry disease: changing pattern of causes of death in FOS - Fabry Outcome Survey. J Med Genet. 2009;46(8):548–52.

6. Namdar M, Kampmann C, Steffel J, Walder D, Holzmeister J, Luscher TF, et al. PQ interval in patients with Fabry disease. Am J Cardiol. 2010;105(5):753–6.

7. Weidemann F, Niemann M, Breunig F, Herrmann S, Beer M, Stork S, et al. Long-term effects of enzyme replacement therapy on fabry cardiomyopathy: evidence for a better outcome with early treatment. Circulation. 2009;119(4):524–9.

8. Hsu TR, Hung SC, Chang FP, Yu WC, Sung SH, Hsu CL, et al. Later Onset Fabry Disease, Cardiac Damage Progress in Silence: Experience With a Highly Prevalent Mutation. J Am Coll Cardiol. 2016;68(23):2554–63.

9. Cortez DL, Schlegel TT. When deriving the spatial QRS-T angle from the 12-lead electrocardiogram, which transform is more Frank: regression or inverse Dower? J Electrocardiol. 2010;43(4):302–9.

10. Kozor R, Callaghan F, Tchan M, Hamilton-Craig C, Figtree GA, Grieve SM. A disproportionate contribution of papillary muscles and trabeculations to total left ventricular mass makes choice of cardiovascular magnetic resonance analysis technique critical in Fabry disease. J Cardiovasc Magn Reson. 2015;17:22.

11. Maceira AM, Prasad SK, Khan M, Pennell DJ. Normalized left ventricular systolic and diastolic function by steady state free precession cardiovascular magnetic resonance. J Cardiovasc Magn Reson. 2006;8(3):417–26.

12. Nordin S, Kozor R, Baig S, Abdel-Gadir A, Medina-Menacho K, Rosmini S, et al. Cardiac Phenotype of Prehypertrophic Fabry Disease. Circ Cardiovasc Imaging. 2018;11(6):e007168.

13. Moon JC, Messroghli DR, Kellman P, Piechnik SK, Robson MD, Ugander M, et al. Myocardial T1 mapping and extracellular volume quantification: a Society for Cardiovascular Magnetic Resonance (SCMR) and CMR Working Group of the European Society of Cardiology consensus statement. J Cardiovasc Magn Reson. 2013;15:92.

14. Messroghli DR, Moon JC, Ferreira VM, Grosse-Wortmann L, He T, Kellman P, et al. Clinical recommendations for cardiovascular magnetic resonance mapping of T1, T2, T2* and extracellular volume: A consensus statement by the Society for Cardiovascular Magnetic Resonance (SCMR) endorsed by the European Association for Cardiovascular Imaging (EACVI). J Cardiovasc Magn Reson. 2017;19(1):75.

15. Vondrak JP, M; Jurek F. . Selected transformation methods snfd their comparison for VCG leads deriving. Alexandria Engineering Journal. 2022;61:3475–85.

16. Potter SL, Holmqvist F, Platonov PG, Steding K, Arheden H, Pahlm O, et al. Detection of hypertrophic cardiomyopathy is improved when using advanced rather than strictly conventional 12-lead electrocardiogram. J Electrocardiol. 2010;43(6):713–8.

17. Schlegel TT, Kulecz WB, Feiveson AH, Greco EC, DePalma JL, Starc V, et al. Accuracy of advanced versus strictly conventional 12-lead ECG for detection and screening of coronary artery disease, left ventricular hypertrophy and left ventricular systolic dysfunction. BMC Cardiovasc Disord. 2010;10:28.

18. Lindow T, Palencia-Lamela I, Schlegel TT, Ugander M. Heart age estimated using explainable advanced electrocardiography. Sci Rep. 2022;12(1):9840.

19. Namdar M. Electrocardiographic Changes and Arrhythmia in Fabry Disease. Front Cardiovasc Med. 2016;3:7.

20. Namdar M, Steffel J, Vidovic M, Brunckhorst CB, Holzmeister J, Luscher TF, et al. Electrocardiographic changes in early recognition of Fabry disease. Heart. 2011;97(6):485–90.

21. Augusto JB, Johner N, Shah D, Nordin S, Knott KD, Rosmini S, et al. The myocardial phenotype of Fabry disease pre-hypertrophy and pre-detectable storage. Eur Heart J Cardiovasc Imaging. 2021;22(7):790–9.

22. Christopoulos G, Graff-Radford J, Lopez CL, Yao X, Attia ZI, Rabinstein AA, et al. Artificial Intelligence-Electrocardiography to Predict Incident Atrial Fibrillation: A Population-Based Study. Circ Arrhythm Electrophysiol. 2020;13(12):e009355.

23. Attia ZI, Noseworthy PA, Lopez-Jimenez F, Asirvatham SJ, Deshmukh AJ, Gersh BJ, et al. An artificial intelligence-enabled ECG algorithm for the identification of patients with atrial fibrillation during sinus rhythm: a retrospective analysis of outcome prediction. Lancet. 2019;394(10201):861–7.

24. Sanz-Garcia A, Cecconi A, Vera A, Camarasaltas JM, Alfonso F, Ortega GJ, et al. Electrocardiographic biomarkers to predict atrial fibrillation in sinus rhythm electrocardiograms. Heart. 2021;107(22):1813–9.

25. Sweda R, Sabti Z, Strebel I, Kozhuharov N, Wussler D, Shrestha S, et al. Diagnostic and prognostic values of the QRS-T angle in patients with suspected acute decompensated heart failure. ESC Heart Fail. 2020;7(4):1817–29.

26. Ladejobi AOM-I, J. R; Shelly Cohen, M; Attia, Z. I; Scott, C; G; LeBrasseur, N. K; Gersh, B. J; Noseworthy, P. A; Friedman, P. A; Kapa, S; Lopez-Jimenez, F. . The 12-lead electrocardiogram as a biomarker of biological age. . European Heart Journal - Digital Health. 2021;2:379–89.

27. Vijapurapu R, Kozor R, Hughes DA, Woolfson P, Jovanovic A, Deegan P, et al. A randomised controlled trial evaluating arrhythmia burden, risk of sudden cardiac death and stroke in patients with Fabry disease: the role of implantable loop recorders (RaILRoAD) compared with current standard practice. Trials. 2019;20(1):314.

